# Socioeconomic status effects on health vary between rural and urban Turkana

**DOI:** 10.1101/2021.07.23.21260771

**Authors:** Amanda J. Lea, Charles Waigwa, Benjamin Muhoya, Francis Lotukoi, Julie Peng, Lucas P. Henry, Varada Abhyankar, Joseph Kamau, Dino Martins, Michael Gurven, Julien F. Ayroles

## Abstract

**Background and objectives:** Understanding the social determinants of health is a major goal in evolutionary biology and human health research. Low socioeconomic status (often operationalized as absolute material wealth) is consistently associated with chronic stress, poor health, and premature death in high income countries. However, the degree to which wealth gradients in health are universal—or are instead made even steeper under contemporary, post-industrial conditions—remains poorly understood.

**Methodology:** We quantified absolute material wealth and several health outcomes among a population of traditional pastoralists, the Turkana of northwest Kenya, who are currently transitioning toward a more urban, market-integrated lifestyle. We assessed whether wealth associations with health differed in subsistence-level versus urban contexts. We also explored the causes and consequences of wealth-health associations by measuring serum cortisol, potential sociobehavioral mediators in early life and adulthood, and adult reproductive success (number of surviving offspring).

**Results:** Higher socioeconomic status and greater material wealth predicts better self-reported health and more offspring in traditional pastoralist Turkana, but worse cardiometabolic health and fewer offspring in urban Turkana. We do not find robust evidence for either direct biological mediators (i.e., cortisol) or indirect sociobehavioral mediators (e.g., adult diet or health behaviors, early life experiences) of wealth-health relationships in either context.

**Conclusions and implications:** While social gradients in health are well-established in humans and animals across a variety of socioecological contexts, we show that the relationship between wealth and health can vary within a single population. Our findings emphasize that changes in economic and societal circumstances may directly alter how, why, and under what conditions socioeconomic status predicts health.

**Lay summary:** High socioeconomic status predicts better health and more offspring in traditional Turkana pastoralists, but worse health and fewer offspring in individuals of the same group living in urban centers. Together, our work shows that under different economic and societal circumstances, wealth effects on health may manifest in very different ways.

## Introduction

A major goal in evolutionary biology and human health research is to understand the social determinants of health, defined as the “distribution of money, power, and resources at global, national, and local levels” that shape health outcomes [1]. Mounting research has shown that these social environmental effects can be profound. In the United States, individuals in the lowest socioeconomic class (defined by absolute material wealth in the form of income) are at greater risk for major health issues such as heart disease, cancer, and diabetes, and are predicted to die over a decade earlier than individuals in the highest socioeconomic class [2–4]. These socioeconomic status (SES) gradients in disease risk and survival are to some degree explained by differences in health care, health habits, and access to resources that are also socially stratified [5]. However, studies in social mammals, where such confounds can be avoided, support the hypothesis that some portion of the SES-health relationship in humans is driven by direct and causal effects of social status on physiology. In particular, animal studies have found that low ordinal dominance rank, a commonly used approximation for low SES in human societies, leads to stress-induced health issues by fundamentally altering hypothalamic-pituitary-adrenal (HPA) axis function [1,6–8].

While there is clear support for the idea that higher SES (operationalized as greater absolute material wealth) is associated with better health in humans, most evidence to date comes from studies of high-income countries (HICs). There is strong appreciation that we need to study the social determinants of health across a wider variety of contexts, and while research in other contexts is rapidly expanding, this body of literature still lags behind what exists for HICs [9–12]. This disparity has made it difficult to comprehensively assess whether the relationship between SES and health is universal and consistent, or instead varies as a function of resource availability and distribution, the nature of social relationships and hierarchies, or other socioecological features of a population (as has been shown for other species [13–15]). In particular, it has been hypothesized that the steep wealth-based gradients in health observed in “Western” HICs are recent byproducts of environmental changes precipitated by urbanization, globalized markets, capitalism, and other modern advancements [16,17]. In other words, while social gradients in health have deep roots in primate and human evolution [1,7], the nature and magnitude of SES-health gradients have potentially changed as a function of modern lifeways.

There are several potential explanations for why post-industrial conditions may exacerbate wealth/SES effects on health. First, relative to small-scale, subsistence-level groups such as hunter-gatherers, modern societies exhibit limited upward mobility and reduced kin support, as well as deep structural racism and violence which may intensify stress in the lowest socioeconomic strata [8,16,18]. Second, modern societies also exhibit a long list of socioeconomically stratified health care resources and health habits (e.g., obesogenic diet, drug and alcohol use) that were largely absent during pre-industrial periods [19–22]. Finally, epidemiological changes that go hand in hand with industrial transitions could alter the nature of SES-health relationships: most deaths in modern day HICs are attributed to non-communicable rather than infectious diseases, and these disease classes are likely differentially affected by wealth. However, because there is a relatively limited literature examining SES effects on health outside of the industrialized setting (e.g., in pre-industrial societies or small-scale, subsistence-level groups), the degree to which urbanization and market-integration fundamentally change the strength or nature of SES-health relationships remains poorly understood [16,23–28].

To address this gap, we quantified the relationship between SES (defined here as absolute material wealth) and health in a small-scale, subsistence-level pastoralist population—the Turkana people of northwest Kenya. Pastoralists are often portrayed as egalitarian, largely because of their relatively equal and open access to natural resources, trading of livestock holdings, and resource sharing during times of hardship [29–31]. However, livestock holdings among pastoralists are also highly correlated across generations, and the intergenerational transmission of wealth inequality is on par with or even greater than what is observed in the most unequal HICs [32] (Gini coefficient estimate for pastoralists [32] versus the US [33]: 0.42±0.05 versus 0.37). Among the Turkana specifically, there is extreme variation in livestock holdings within and between generations, crossing several orders of magnitude and often fluctuating due to unpredictable events such as droughts, livestock disease outbreaks, or raiding from nearby groups [30]. This complex picture of high inequality and variance in absolute wealth paired with egalitarian practices makes it unclear to what degree we should expect SES to affect health outcomes in pastoralist societies like the Turkana. While previous work with pastoralist communities has examined effects of herd size (the primary source of material wealth) on nutrition and caloric intake [34–36], little work has tested the relationship between SES and health in traditional pastoralist societies [35,37,38].

Our study set out to explore SES-health connections in traditional Turkana, and to ask how these links strengthen or change when individuals transition to a more urban, market-integrated lifestyle. We were able to perform this comparison because cultural and economic changes paired with expansion of country-wide infrastructure has prompted many Turkana to move to densely populated cities over the last few decades. Turkana migrants to urban areas no longer practice pastoralism, work wage labor or market-interfacing jobs, and experience many other lifestyle changes. For example, urban-dwelling Turkana consume fewer traditional and more processed foods relative to pastoralist Turkana, which puts them at greater risk of cardiometabolic disease [39]. Moving to urban areas may also lead to increased psychosocial stress, reduced kin support, and changes in health habits and physical activity—all of which impact health and may vary with SES [7,40,41].

By collecting data on absolute material wealth and health from both traditional, pastoralist and urban, market-integrated Turkana, we were able to test whether lifestyle change alters SES effects on health within a single population. We also performed three sets of follow up analyses to understand the causes and consequences of SES-health associations in both rural and urban contexts. First, to understand the putative fitness consequences of SES, we tested for SES effects on reproductive success (number of surviving offspring). SES consistently predicts reproductive success in natural fertility contexts where wealth is largely somatic/embodied or relational, such that extra-somatic material wealth can be used to enhance reproduction [42,43]. However, in socioeconomic landscapes where greater human capital investment is needed to compete successfully in labor markets, SES is often decoupled from reproduction. In this post-“fertility transition” context, which we speculate at least partially reflects our urban sample, the relationship between SES and health is often less clear and worth exploring. Second, to understand the biological and behavioral mediators of SES-health connections, we measured 1) serum cortisol to assess the role of psychosocial stress and HPA axis function and 2) we interviewed study participants about their diet, health habits, and use of health care resources. We then asked whether SES predicted any of these potential mediators, and if so, we performed formal mediation analyses to understand the proportion of the total effect that was mediated [44,45]. Finally, we were interested in understanding another key social determinant of health— early life adversity (ELA)—which has been shown to both affect later life health outcomes [1,46,47] and to set individuals on a course toward low SES in adulthood [47–49]. We therefore devoted effort toward systematically documenting variation in early life experiences and exploring their effects, which has rarely been attempted in small-scale, subsistence-level groups [50–52]. Taken together, our study provides a comprehensive picture of how, why, and under what circumstances SES affects health. We leverage the lifestyle gradient of the Turkana to directly address the impact of increasing urbanization and market-integration on this important relationship.

## Methods

### Overview of the study population and study methodology

The Turkana have resided in northwest Kenya since the early 18th century [53]; their homelands (Turkana county) are semi-arid and characterized by low annual rainfall, frequent droughts, and high year round temperatures [54]. The Turkana people are traditionally nomadic pastoralists, relying on dromedary camels, zebu cattle, fat tailed sheep, goats, and donkeys for subsistence [55]. Most herders keep livestock from all species, though they may specialize to some degree [31]. As a result of their subsistence strategy, the traditional Turkana diet is extremely protein-rich: 70-80% of calories are derived from milk or other animal products [55]. For detailed descriptions of the diet, climate, and lifestyle experienced by traditional, pastoralist Turkana, see work from the South Turkana Ecosystem Project [56].

Over the last several decades, many urban areas in central Kenya have experienced an influx of Turkana people as a result of country-wide infrastructure improvements and rapid cultural, economic, and social changes; for the same reasons, the capital of Turkana county (Lodwar) has also become increasingly urban and market-integrated. In our study, we use the term “urban” to refer to people living in densely populated cities characterized by many permanent businesses, and where most people engage with the market economy and/or work wage labor jobs. We defined “urban” individuals as those that no longer practice pastoralism and reside in one of three cites included in our study—Nanyuki, Lodwar, and Kitale. All three of these cities have population sizes >20k and are among the top 50 largest cities in Kenya (https://worldpopulationreview.com/). We also included non-pastoralists residing in suburbs in Laikipia county in central Kenya in the urban category, because Laikipia is a cosmopolitan area with several large cities (e.g., Nanyuki, Nyahururu, and Rumuruti). We note that individuals that choose to move to urban areas likely represent a non-random subsample of the Turkana population; we do not currently have data on the economic or social considerations that motivate individuals to migrate, though this is a focus of ongoing work.

We defined pastoralists as residents of Turkana county who self-reported their main subsistence activity as “pastoralism”, who owned livestock, and who drink milk every day (i.e., they rely on their livestock for subsistence). In previous work, we also defined a third category of Turkana who no longer practice pastoralism but still live in the relatively remote and rural Turkana homelands [39]. For the purposes of this study, we focused only on the extremes of the Turkana lifestyle spectrum (pastoralist versus urban) because SES is more difficult to define and operationalize in the intermediate context, where both livestock and material goods contribute to absolute material wealth (see next section).

Data were collected in Turkana, Laikipia, and Trans-Nzoia counties between April 2018 and February 2020. During this time, researchers visited locations where Turkana individuals were known to reside (Figure S1). At each sampling location, local chiefs and elders were first consulted about the project. If they believed the study to be of interest to their community, a larger meeting was held to explain the project to all interested individuals. After this period of discussion, adults (>18 years old) of self-reported Turkana ancestry were invited to participate in the study. The study involved a structured interview, blood sample collection, and anthropometric measurements. Additional background on the study as well as detailed sampling procedures and demographic summaries are provided in [39].

### Structured interviews

#### Self-reported health, potential behavioral mediators, and covariates

Structured interviews were conducted with all participants to collect information about demography, reproductive history, diet, early life experiences, lifestyle, and absolute material wealth. All interviews were conducted in a language familiar to the participant (English, Turkana, or Swahili). The following self-reported variables from the interviews are relevant to our analyses:

- Sex
- Age
- Main subsistence activity, chosen from the following categories: self-employment, formal employment, petty trade, farming, pastoralism, hunting and gathering, other
- Highest education level, scored as: 0=none, 1=lower primary school, 2=upper primary school, 3=secondary school, 4=education beyond secondary school
- Number of surviving children
- Number of wives (for men only)
- Whether the participant used contraceptives (for women only; Y/N)
- Whether the participant used medications or sought medical treatment when ill in the last month (Y/N/NA, if not ill in the last month)
- Whether the participant used alcohol, tobacco, or cigarettes (never/occasionally/daily)
- Whether the participant was currently fasting (this covariate was included in analyses of blood glucose)
- Whether the participant experienced each of the following health issues in the last 3 months (Y/N for each question): swollen extremities, fatigue or weakness, shortness of breath, diarrhea, worm infection, stomach pain, vomiting, constipation, coughing, difficulty breathing, dizziness, headaches, chest pain, bruises, cuts and scrapes, or burning during urination.

We also used a food frequency questionnaire to collect information about the consumption of meat, milk, bread, sugar, salt, and cooking oil. We focused on these items because they reflect foods that are essential (meat, milk) or uncommon (bread, sugar, salt, cooking oil) in the diet of traditional pastoralists. Participants were asked how often a specific item was used or consumed and were given the following answer choices: never, rarely, 1-2 times per week, >2 times per week, or every day. These answers were converted to a scale of 0-4. Results for sugar, salt, and cooking oil were then tallied and combined into a single metric because these answers were highly correlated (all R^2^>0.9).

Because 1) we were interested in a life course perspective on the social determinants of health and 2) early life challenges can set individuals on a course toward adverse socioeconomic circumstances in adulthood [47–49], we interviewed participants about their early life experiences, adapting the CDC’s Adverse Childhood Experiences (ACEs) instrument [57]. While this instrument has not been previously used with the Turkana, it has been applied in hundreds of studies [58–60], including in low- and middle-income settings and in Kenya specifically [61–63]. We created a tally of the number of adversities each participant experienced as a child (<12 years old) from the following list: mother or father absent (i.e., didn’t leave with the participant when they were young due to death, abandonment, or other circumstances), verbal abuse or threat of violence by a caregiver, physical abuse by a caregiver, witness of verbal or physical abuse toward mother, exposure to mental illness, exposure to alcoholism or other substance abuse, and food insecurity. These questions were asked using the phrasing provided by the Centers for Disease Control (https://www.cdc.gov/violenceprevention/aces/ace-brfss.html) (modeled after [57]), except for the food insecurity question, which we added because it is a common concern in Kenya. We also asked participants where they were born, as well as what the main subsistence strategy and occupation of their parents were during childhood.

#### SES (absolute material wealth) metrics

We drew on our interview data to create two separate metrics of SES, one for the pastoralist context and for the urban context. Our measures are meant to capture key indicators of absolute material wealth in each context, rather than other social determinants of health such as influence or standing in the community (as perceived by the focal individual or other community members), embodied wealth, relational wealth, or inequality/relative material wealth. We focused on absolute material wealth because this particular social determinant of health is well-established in the literature and often has the strongest effect sizes [64,65].

Livestock are the primary source of material wealth among pastoralists [29]. Therefore, in the pastoralist setting, we defined SES as the total number of multispecies livestock owned by the household the individual belonged to, following [29] and references therein. This value was log_2_ transformed because of skew. This measure of SES was strongly correlated with other possible metrics of material wealth [35,66], for example livestock translated into estimated market prices (β=1.01, p-value<10^−16^, linear regression) as well as the ratio of total multispecies livestock holdings to the number of household members (β=0.913, p-value<10^−16^, linear regression).

In the urban setting, we used a tally of durables/goods, dwelling characteristics, and other household assets as an index of SES and absolute material wealth. This approach is relatively common [67] and the specific list was derived from previous work [68–70]. We tallied household possession of the following items to create an index ranging from 0-11: finished floor, finished roof, electricity, television set, mobile phone, flush toilet, gas cooking, indoor tap water, treated water, >1 room in the household, and <= 2 household members per room. We did not include durables and dwelling characteristics in the pastoralist SES index, because very few individuals in the pastoralist setting own any of goods listed above (3.1%). Similarly, we did not include livestock holdings in our urban SES metric, because few individuals in this setting own livestock (21.4%). Further, livestock holdings are strongly correlated with the urban SES index, suggesting we are not missing primary sources of wealth by not counting livestock (β=0.027, p-value=0.036, Poisson regression). Our urban SES index is also strongly associated with education levels (β=0.203, p-value<10^−16^, Poisson regression), another common measure of SES in HICs and industrialized settings [71].

Defining absolute material wealth is a complex and challenging task, and many approaches have been taken [67,72]. The approach described above is based on data that were feasible to collect, and precedent in the literature; however, one drawback is that, by necessity, our measures of SES are different in the pastoralist versus urban context and therefore not directly comparable. We believe this is appropriate given the strong socioeconomic differences between the pastoralist and urban contexts. However, to understand the robustness of our results, we also performed supplementary analyses in which we 1) applied the urban or pastoralist measures of SES described above to both contexts or 2) performed principal components analysis of data on livestock holdings, educational attainment, and household assets to create a single measure of SES that we then applied to both contexts. The results of these supplementary analyses agree with the direction of effects and overall conclusions presented in the main text, and are described in greater detail in the Supplementary Materials.

### Measuring biomarkers of cardiometabolic health

In addition to collecting data on self-reported health, we also measured 10 biomarkers of cardiometabolic health. We used standard anthropometric approaches to measure body mass index (BMI), waist circumference, body fat percentage, as well as systolic and diastolic blood pressure. We also collected venous blood to measure blood glucose levels, total cholesterol, triglycerides, and high- and low-density lipoproteins (HDL, LDL). The collection of all of these measures are described in detail in [39] and are also repeated in the Supplementary Materials.

Data were excluded in a handful of cases where individuals were extreme outliers, indicating a likely error (>5 standard deviations from the mean). Prior to statistical analyses, all biomarkers and SES measures (when they were used as predictor variables) were mean-centered and scaled by their standard deviation using the “scale” function in R [73]. Consequently, all effect sizes reported in the methods are standardized, and represent the effect of a given variable on the outcome in terms of increases in standard deviations.

### Measuring serum cortisol

As a proxy for HPA axis function, we measured serum cortisol. Cortisol is known as a “stress” hormone and is released in response to physical and or psychosocial sources of stress. However, cortisol also has other biological functions, for example it is involved in blood pressure maintenance, immune function, and both protein and carbohydrate metabolism [74]. Thus, while we focus on cortisol as a measure of HPA axis function, we note that variation in this hormone could also be driven by SES effects on other biological processes.

To collect serum, venous blood was drawn from each participant into a serum separator tube (Fisher Scientific) and spun immediately for 15 minutes at 2500 RPM in a portable centrifuge. The serum layer was then pulled off the top of the tube, transferred to a 2mL cryovial, and frozen at −10C in a portable freezer. Samples were kept in the portable freezer for no longer than one week, after which they were transferred to long-term storage in a −20C freezer at Mpala Research Centre (Laikipia, Kenya). Samples were exported to the United States on dry ice, and upon arrival were stored at −80C.

216 serum samples were thawed on ice and used to quantify cortisol with the Cortisol Elisa Assay Kit from Eagle BioSciences, according to the manufacturer’s instructions. Samples were randomized across three plates, and the R^2^ between the expected and observed concentrations for the calibrator curves were 0.9994, 0.9989, and 0.9977. Two control samples of known concentration were run on each plate; all control sample values fell within the acceptable range specified by the kit’s manufacturer, and the between-plate coefficient of variation for these two samples was 0.0326 and 0.0391, respectively. In the total sample, we did not find any evidence for plate effects (ANOVA, F=1.328, p-value=0.267, n=216) and we did observe expected effects of age, sex, and time of day of sample collection [75–77]. Specifically, our linear models revealed that males exhibited higher cortisol levels than females (β=-0.484, p-value=0.013), cortisol levels increased with age (β=0.021, p-value=0.006), and cortisol levels were higher in the morning (β=-0.104, p-value=0.018). In a linear model controlling for the above covariates, we did not find evidence of mean differences in cortisol levels between pastoralist and urban individuals (β=-0.139, p-value=0.489).

### Testing for SES effects on health in the pastoralist and urban settings, and exploring potential mediators

For each of the 16 self-reported health measures, we used binomial regression to ask whether SES was predictive of health in each context. For each of the 10 biomarkers of cardiometabolic health, we used linear models to ask whether SES was predictive of health outcomes in the urban or pastoralist setting (analyzed separately; see Table S1 for sample sizes). We did not run models in cases where <1% of people experienced a given self-reported health issue (Table S1). All analyses controlled for self-reported age and sex as fixed effects covariates. For each health outcome, we also ran analyses that considered a sex x SES interaction term, and we used the results of the second model if the delta AIC between this model and the first model was >2 (Table S1). All p-values were corrected for multiple hypothesis testing using a Benjamini-Hochberg false discovery rate approach [78]. We considered SES effects to be significant in a given context at a 10% FDR cutoff.

We also explored potential mediators of SES-health associations. To do so, we used formal mediation analyses [44,45] following the methods in [6,39,79]. We considered the following behavioral factors as potential mediators: cigarette smoking (Y/N), alcohol usage (Y/N), tobacco usage (Y/N), use of health care resources (i.e., whether the participant used medications or sought medical treatment when ill; Y/N/NA), frequency of use of salt/sugar/oil in cooking (0-4), and frequency of consumption of bread, meat, or milk (0-4). We also considered cortisol levels as a potential biological mediator. For a variable to be a potential mediator, it needs to be correlated with the predictor variable of interest. We therefore used linear, binomial, and Poisson regression (for continuous, binary, and count variables, respectively) to predict each mediator as a function of SES, in the pastoralist and urban samples, respectively. All models controlled for age and sex, and the cortisol models also controlled for time of day of sample collection.

We tested all variables for mediation that were predicted by SES at a relaxed nominal p-value of 0.1, and that were associated with SES in a direction that made sense for mediating health effects. To do so, we fit two models: 1) an “unadjusted” model that included the effect of the predictor variable on the outcome (i.e., the effect of SES on a given health outcome, controlling for covariates) and 2) an “adjusted” model that is identical to model 1 but also includes the putative mediator as a covariate. If the predictor’s effect on the outcome is explained by the mediator, then the effect of the predictor (β SES) will decrease when the mediator is included in the adjusted model and absorbs variance otherwise attributed to the predictor. To assess significance, we estimated the decrease in β SES between the unadjusted and adjusted models across 1000 iterations of bootstrap resampling. We considered a variable to be a significant mediator if the lower bound of the 95% confidence interval for the decrease in β SES did not overlap with 0.

### Testing for SES effects on reproductive success in the pastoralist and urban settings

We were also interested in the potential fertility consequences of SES, as identifying these links is important for thinking about how SES effects on health may ultimately impact Darwinian fitness, and thus for informing our understanding about how SES-associated traits (e.g., striving for wealth or other forms of status) evolve in humans [42]. Therefore, we used Poisson regression to predict the number of surviving offspring each individual had as a function of SES, age, sex, and the interaction between SES and sex (as this improved model fit in both contexts). The total number of surviving offspring was self-reported, and was not available in terms of offspring sex or age breakdowns (e.g., we could not calculate number of offspring surviving past a certain age). We also note that number of surviving offspring is not the same as Darwinian fitness, which is difficult to measure, but it is a commonly used proxy in the literature and is routinely considered a fitness-related trait (e.g., [42]).

For men in the pastoralist setting, we ran a post hoc analysis to understand whether SES effects on reproductive success were mediated by SES effects on number of wives. In traditional Turkana culture, livestock are used as bridewealth and polygyny is common [31]. This practice is much less common in urban settings: 42% versus 9% of married men reported >1 wife in the pastoralist and urban settings, respectively. To test whether SES effects on reproductive success were mediated by SES effects on number of wives, we fit two Poisson regression models to data from male pastoralists: 1) an “unadjusted” model that included the effect of SES on number of children, controlling for age and 2) an “adjusted” model that was identical to model 1 but also included number of wives as a covariate. We used the same bootstrap approach described above to assess significance.

For women in the urban setting, we ran a post-hoc analysis to understand whether SES effects on reproductive success were mediated by SES effects on birth control usage. Contraceptive use is common among urban women but highly uncommon among pastoralist women: 51% versus 0.6%, respectively. To test birth control usage as a potential mediator, we again fit an “unadjusted” model and an “adjusted” model (that included birth control usage as a covariate) and used a bootstrap approach to assess significance.

### Understanding the relationships between lifestyle, early life adversity, adult SES, and adult health

To test whether early life adversity predicted adult SES in each context, we used linear regression (for pastoralists) and Poisson regression (for urban individuals) to predict each wealth index as a function of our cumulative ELA score controlling for age and sex. Cumulative ELA was considered on a scale of 0-5, with individuals experiencing >5 adversities collapsed into the 5 category to prevent the influence of outliers. We also used Poisson regression to understand whether cumulative ELA itself was predicted by 1) age, sex, or lifestyle (urban versus pastoralist) in the total sample and 2) age, sex, or parental subsistence strategy (coded as pastoralist, other, or formal employment) in the pastoralist sample only.

To test whether ELA was related to adult health in each context, we used linear regression controlling for age and sex to predict our 10 biomarkers of cardiometabolic health as a function of cumulative ELA. Unfortunately, we did not conduct interviews on self-reported health and early life experiences for the same set of individuals, and we therefore could not assess the relationships between these measures. All p-values were corrected for multiple hypothesis testing and were considered significant if they passed a 10% FDR.

### Ethics approval

This study was approved by Princeton University’s Institutional Review Board for Human Subjects Research (IRB# 10237), and Maseno University’s Ethics Review Committee (MSU/DRPI/MUERC/00519/18). We also received county-level approval for research activities, and research permits from Kenya’s National Commission for Science, Technology and Innovation (NACOSTI/P/18/46195/24671). Written, informed consent was obtained from all participants after the study goals, sampling procedures, and potential risks were discussed with community elders and explained to participants in their preferred language (by both a local official, usually the village chief, and by researchers or field assistants).

## Results

### Social status effects on health and a fitness-related trait are highly context-dependent

Among traditional pastoralists, SES was positively associated with self-reported health. High status individuals were significantly less likely to suffer from recent chest pain (β SES=-0.624, q-value=0.016), diarrhea (β SES=-1.472, q-value=0.057), vomiting (β SES=-2.604, q-value=0.074), dizziness (β SES=-1.424, q-value=0.016), and fatigue or weakness (β SES=-1.094, q-value=0.016; Table S1-3). These effect sizes were substantial: for example, 33%, 14%, and 9% of individuals in the lowest quartile of the SES distribution experienced chest pain, fatigue/weakness, and dizziness in the past three months, compared to 8%, 7%, and 0% of individuals in the highest quartile (Figure 1B). There were no significant relationships between SES and measures of cardiometabolic health among traditional pastoralists (Table S1); most pastoralists were cardiometabolically healthy (Figure S2).

**Figure 1.**
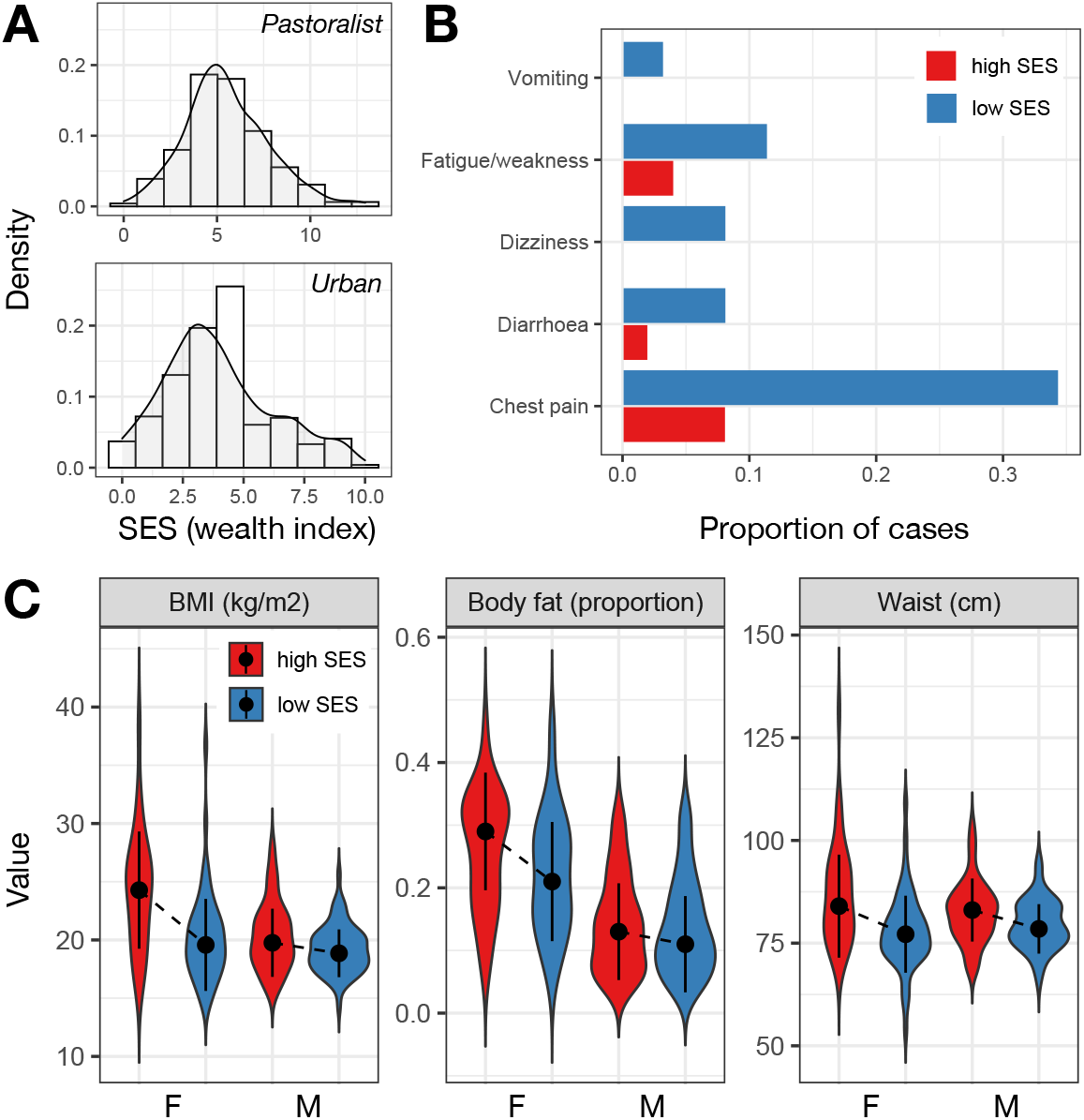
Social status effects on health are context-dependent, and vary by sex and lifestyle. (A) Distribution of SES measures among pastoralists (where SES was defined as log2 transformed livestock counts) and urban individuals (where SES was defined by a tally of market-derived possessions). (B) Proportion of people reporting various health issues as a function of SES in the pastoralist setting. For visualization, data from individuals in the highest and lowest SES quartiles are plotted. (C) Distribution of cardiometabolic biomarker values as a function of sex and SES (highest versus lowest quartiles) in the urban setting. Dots represent the median of each distribution and solid lines represent the median +/- 1 standard deviation.

The direction of the effect of SES on health was reversed in the urban setting and captured entirely different health outcomes. In this context, we found no relationships between SES and self-reported health, even though the frequency of self-reported health issues was higher than in the pastoralist setting (Table S1). Among urban individuals, high status was associated with biomarkers of worse cardiometabolic health: higher BMI (β SES=0.425, q-value=1.02×10^−7^), waist circumference (β SES=0.436, q-value=7.46×10^−9^), diastolic blood pressure (β SES=0.113, q-value=0.088), and body fat (β SES=0.215, q-value=5.82×10^−4^; Table S1-3). With respect to BMI, a modest number of individuals in the urban setting met the criteria for being overweight (12.56%) or obese (2.90%); nevertheless, BMI is often a linear predictor of type II diabetes [80] and cardiovascular disease [81] (though not necessarily of mortality [82]), suggesting that this SES-driven variation is still potentially meaningful. For all of the above traits except blood pressure, we observed that women were more sensitive to SES effects on health (body fat: β SES x sex=-0.157, p-value=0.03; waist circumference: β SES x sex=-0.225, p-value=0.012; BMI: β SES x sex=-0.313, p-value=8.63×10^−4^; Figure 1C).

In both the pastoralist and urban settings, SES was associated with reproductive success, but in opposite directions. In the urban setting, high SES was associated with fewer surviving offspring (β SES= −0.129, p-value=8.68×10^−4^), and an interaction effect pointed to this effect being primarily driven by stronger effects in women than men (β SES x sex=0.136, p-value=0.010; Figure 2 and Table S4). Causal mediation analyses revealed that the effect of urban female SES on reproductive success is partially explained by SES effects on contraceptive usage (estimated proportion of the total effect that is mediated=12.6%, p-value=0.020). In the pastoralist setting, a main effect of SES was not significant (β SES=-6.86×10^−3^, p-value=0.846), but an interaction term supporting stronger SES effects in men than women was significant (β SES x sex=0.095, p-value=0.047; Figure 2 and Table S4). Mediation analyses revealed that the effect of male pastoralist SES on reproductive success is partially explained by SES effects on the number of wives a man had (estimated proportion of the total effect that is mediated=25%, p-value=0.046; Figure S3).

**Figure 2.**
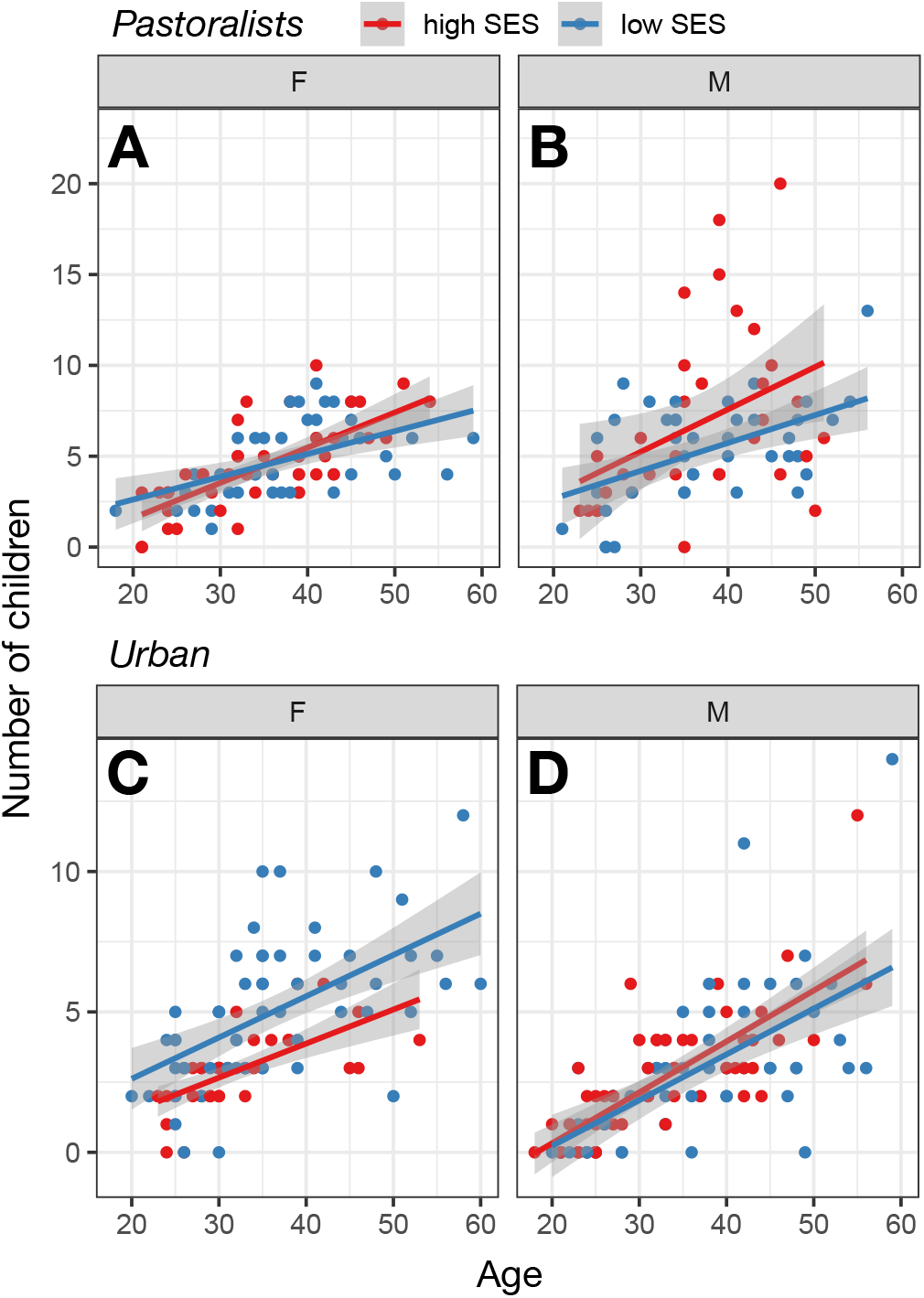
Social status effects on number of surviving offspring are context-dependent, and vary by sex and lifestyle. (A-B) Number of living children as a function of age among pastoralist women and men in the highest versus lowest SES quartiles. (C-D) Number of living children as a function of age among urban women and men in the highest versus lowest SES quartiles.

### Mediators of the SES-health relationship among pastoralist and urban Turkana

We explored the role of cortisol as well as sociobehavioral variables in mediating links between SES and health. We did not find any association between SES and serum cortisol in either the pastoralist (β SES=0.103, p-value=0.577, N=79) or urban samples (β SES=0.025, p-value=0.887, N=59) (Figure S4). Therefore, we did not test this variable for mediation. In terms of sociobehavioral variables, several of the factors we hypothesized might explain social gradients in health were also not predicted by SES, and were therefore excluded from further mediation analyses. In particular, SES was unrelated to the usage of cigarettes, alcohol, tobacco, health care resources, meat, and bread in the pastoralist setting (Table S5). In the urban setting, SES was unrelated to usage of cigarettes and health care resources (Table S5-6). SES was associated with alcohol (β SES=-0.254, p-value=0.019) and tobacco usage (β SES=-0.658, p-value=3.56×10^−4^) among urban Turkana, but the direction of these effects was not compatible with mediation (i.e., low SES individuals exhibited worse health habits).

Because they were significantly or marginally associated with SES, we tested whether the following variables could explain observed SES-health associations: 1) greater usage of salt, sugar, and oil among low SES pastoralists (β SES=-1.304, p-value<2×10^−16^) and 2) greater consumption of meat (β SES=0.150, p-value=1.29×10^−5^), bread (β SES=0.150, p-value=2.18×10^−13^), and milk (β SES=0.141, p-value=3.13×10^−5^), as well as greater reliance on salt, sugar, and oil (β SES=0.026, p-value=0.097), among high SES individuals in the urban setting (Table S5). We found generally minimal evidence for mediation (Table S7), with the exception that salt, sugar, and oil consumption explained an estimated 11% of the effect of SES on waist circumference in urban individuals.

### Lifestyle patterns early life experiences, but early life adversity does not predict adult SES or adult health

Turkana practicing pastoralism as adults experienced greater cumulative early life (ELA) relative to those living in urban settings in adulthood (β lifestyle=-0.465, p-value=1.97×10^−3^; Table S8). We also found that a lifestyle x sex interaction improved model fit, with the direction of this effect suggesting that pastoralist men experienced the highest levels of cumulative ELA (β lifestyle x sex=-0.343, p-value=0.075; Figure 3). For example, 11.1% and 15.6% of pastoralist women and men experienced 5 or more adversities, while 6.3% and 2.9% of urban women and men experienced the same level of hardship. For reference, these numbers are estimated at 10.3% and 6.9% for women and men in the United States [83] (though we note the US numbers are derived from a slightly different questionnaire; Table S9 and Figure S5). Age of the study participant also trended toward a positive association with cumulative ELA in both contexts, suggesting that incidence of ELA has generally reduced over time (β age=0.007, p-value=0.058).

**Figure 3.**
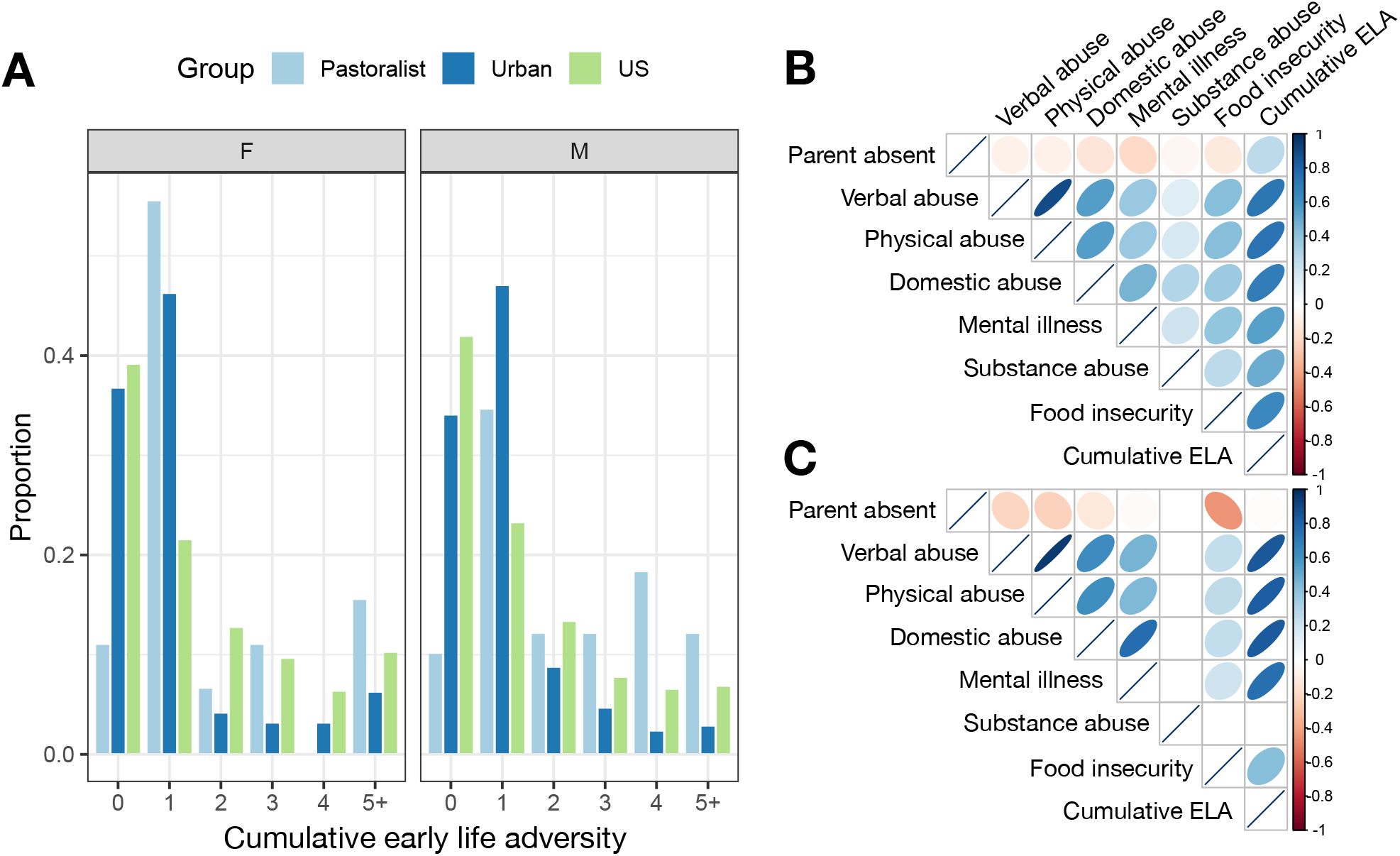
Lifestyle effects on patterns of early life adversity. (A) Number of early life adversities experienced by sex and population. US data were sourced from [83]. Correlations among individual sources of early life adversity as well as cumulative early life adversity tallies for (B) urban and (C) pastoralist individuals. Note that no pastoralist individuals were exposed to substance abuse within their household growing up, and no pairwise correlations are therefore presented that involve this measure.

While individuals practicing pastoralism as adults all grew up in pastoralist families, 46% of urban adults grew up in pastoralist families; the remaining 30% and 24% had parents that relied on formal employment or other types of employment, respectively. We found that, within urban adults specifically, early life subsistence strategy mattered for ELA in ways that were consistent with results from the total sample: individuals who grew up in pastoralist families experienced more ELA than individuals whose parents were formally employed (β parental occupation=-0.275, p-value=0.036). We did not find any significant differences for the other contrasts (i.e., formal employment versus other employment or pastoralism versus other employment, both p>0.05).

While cumulative ELA is clearly patterned by lifestyle, we did not find any compelling evidence that this variation matters for adult health in our sample (Table S10). Controlling for age and sex, we also did not find that cumulative ELA was predictive of adult SES in the urban setting (β ELA=0.031, p-value=0.219). In the pastoralist setting, we observed a trend toward greater cumulative ELA predicting higher adult social status (β ELA=0.268, p-value=0.061). The direction of this marginal effect was unexpected and suggests that cumulative ELA does not mediate the (positive) effects of SES on health we observe in traditional pastoralists.

## Discussion

Here, we test the hypothesis that shifts toward more urban lifestyles alter or exacerbate the relationship between SES and health, by examining SES-health associations within a single group undergoing rapid lifestyle change. Previous work has suggested that the effects of socioeconomic status on health may be exacerbated in HICs relative to ancestral and pre-industrial human societies [16]. In particular, the social conditions under which humans evolved were thought to feature less rigid hierarchies, greater kin support, and greater upward mobility than what is currently observed in HICs [8,16,18]. They also did not routinely feature socioeconomically stratified health care resources, structural racism, and other forms of institutionalized inequality [19–22]. Thus, the negative effects of limited material wealth that are common in HICs could be a recent byproduct of post-industrial societal, economic, or even epidemiological changes (e.g., most deaths in HICs are driven by non-communicable rather than infectious diseases, which could have different relationships to SES). Identifying the degree to which this hypothesis is true is important for understanding the evolution of the social determinants of health as well as the best strategies for reducing health disparities.

We found strong evidence that the recent transition from pastoralism to an urban, market-based lifestyle alters the relationship between absolute material wealth and health among the Turkana. Among traditional Turkana pastoralists, high SES is associated with better self-reported health, but unassociated with cardiometabolic health. In contrast, among urban Turkana, high SES is associated with worse cardiometabolic health. At first glance, these results do not necessarily adhere to the framework laid above, which would predict that low SES would be associated with worse health in both the pastoralist and urban setting, with these effects magnified in the urban setting. Instead, we find that the presence and direction of SES effects are modified as a function of lifestyle. We also found similar opposing effects when examining a fitness-related trait: high SES predicts more offspring in the pastoralist setting (partially driven by wealth effects on polygyny), but fewer offspring in the urban setting. In the urban setting, wealthier women were more likely to use contraceptives and this mediated some of the effect of SES on reproductive success. We speculate that, in the urban setting, wealth may also be associated with later marriage as well as other forms of family planning [84].

While some of our results run counter to the large literature linking low SES to worse outcomes, they highlight the importance of social, ecological, and economic context. In the pastoralist setting, wealth is likely channeled into traditional foods, family growth, and other resources with positive effects on self-reported health. In this setting, it is also possible that high SES individuals are better able to cope with ecological hardship, such as frequent droughts (though we note that follow-up analyses testing for SES x season [85] effects on health in the pastoralist setting did not reveal significant effects; Table S11). In contrast, in the urban setting, wealth is likely channeled into consumption, purchase, and amenities that negatively impact cardiometabolic health (e.g., processed foods, vehicles to minimize physical activity). We found some support for this idea in our mediation analyses, where increased consumption of market-derived foods—namely, salt, sugar, and oil—mediated 11% of the effect of SES on waist circumference in the urban setting. We note that the idea that high socioeconomic status in newly urban Turkana translates into purchases that negatively impact cardiometabolic health (e.g., processed foods) is also consistent with literature on epidemiological and nutritional transitions, in which newly developed countries often suffer an increased burden of non-communicable diseases [86].

The highly context-dependent nature of SES effects observed in the Turkana dovetails with recent work in non-human primates, which has also found that both the magnitude and direction of social status effects varies across systems. For example, previous work has emphasized the association between both low SES in humans and low ordinal dominance rank in rhesus macaques (a well-established model of human SES [8]) and increased expression of innate immune and inflammation-related genes [6,87,88]. However, in wild baboons, weak effects in the expected direction were observed in females [15], while strong patterns in the *opposite* direction were observed in males [79] (i.e., high dominance rank predicted increased expression of inflammation-related genes). Importantly, dominance rank is attained through direct physical competition in male baboons but not in any of the other species or sexes, suggesting that the heterogeneity between studies reflects differences in the nature of social hierarchies. Such a nuanced picture is consistent with decades of work on non-human primates that has emphasized the diverse ways in which social status is attained and maintained across systems, as well as the diverse sets of costs and benefits that are associated with status [7,8]. In these systems, it appears that the physiological and health correlates of dominance rank vary as a function of the specifics of the social environment as well. Our results suggest that the same logic could be applied toward thinking about heterogeneity in SES gradients in health in human societies.

In addition to showing that transitions to urban, market-integrated lifestyles alter the relationship between absolute material wealth and health in a single population, our study also reports novel findings related to early life experiences. In particular, we used the adverse childhood experiences framework to document variation in early life adversity as a function of lifestyle. We found greater ELA in the traditional, pastoralist setting, but no relationship between ELA and adult SES or health in either setting. The lack of an association with health is noteworthy given the large body of literature linking greater ACE exposure to earlier death and later life cardiovascular, autoimmune, and neurodegenerative diseases in HICs [57,89–96]. However, there has been less work on the subject in low- and middle-income countries (but see [61,97]), despite childhood adversity being a growing area of international interest [98]. It may be that larger sample sizes are needed to uncover effects in the Turkana, that early life adversity does not carry the same psychosocial and physiological costs as in HICs because of differing cultural norms [99], or that the ACE questionnaire does not capture the most salient types of adversity experienced by the Turkana (e.g., experiences of livestock loss or raiding in the pastoralist context [38,99,100]). Additionally, retrospectively collected information about childhood experiences may be biased or incomplete [101], and our analyses of ELA effects on self-reported health could suffer from common source bias (though this is not true for our analyses of cardiometabolic biomarkers) [102]. In general, more research is needed that considers a wider variety of social, economic, and cultural circumstances in the study of early life adversity, and that incorporates longitudinal study designs whenever possible to alleviate concerns about retrospective reporting and common source bias.

There are several limitations to the present study, as well as open directions for future work. First, our analyses of SES effects on serum cortisol levels did not reveal any significant associations and were limited by a small sample size. Previous work in human and non-human primates has shown that individuals with low SES or low dominance rank are often chronically stressed, which leads to altered HPA axis function reflected in chronically elevated cortisol levels [1,7,8]. However, social status effects on cortisol are also known to be context-dependent across non-human primate hierarchies [103], suggesting that this may be another area where lifestyle modifies the direction or magnitude of SES effects. Future work could expand the sample set and further explore the context-dependency of cortisol levels across lifestyle groups. Another limitation of the current study is that we were unable to identify sociobehavioral mediators of the SES-health relationship in either the pastoralist or urban setting. It may be that these relationships are not mediated by indirect effects of behavior, and are instead entirely explained by direct effects of SES on biological mechanisms we have yet to uncover (e.g., cortisol levels) or explore (e.g., gene regulation [6,79,87,104]). Alternatively, SES effects may be mediated by behavioral variables that were not captured by our surveys. For example, the relationship between SES, stress, and health may critically depend on the availability of kin and social support [1,30,105], which we did not measure here and which likely varies dramatically as a function of lifestyle.

## Data Availability

Anonymized data underlying the main results will be publicly available on Dryad following publication.

## Acknowledgments

We thank previous members of the Turkana Health and Genomics Project for their contributions, especially S. Lowasa, D. Mukhongo, S. Ngatia, E. Loowoth, and M. Ndegwa. We thank J. Orina, K. Tombak, and D. Rubenstein for logistical help in Kenya and members of the Graham lab at Princeton for technical assistance with the cortisol Elisa assay. We are also grateful to the staff of Mpala Research Centre for their essential support, especially F. Hassan, C. Nzomo, B. Wanjohi, G. Chege, T. Maina, and J. Nakolonyo. Finally, we thank all of the Turkana communities for their contributions to and support of our scientific work.

## References

1. Snyder-Mackler N, Burger JR, Gaydosh L et al. Social determinants of health and survival in humans and other animals. Science (80-) 2020;368, DOI: 10.1126/science.aax9553.

2. Chetty R, Stepner M, Abraham S et al. The Association Between Income and Life Expectancy in the United States, 2001-2014. J Am Med Assoc 2016;315:1750.

3. Stringhini S, Carmeli C, Jokela M et al. Socioeconomic status and the 25 × 25 risk factors as determinants of premature mortality: a multicohort study and meta-analysis of 1·7 million men and women. Lancet 2017;389:1229–37.

4. Marmot M. Public Health Social determinants of health inequalities. 2005;365:1099–104.

5. Kröger H, Pakpahan E, Hoffmann R. What causes health inequality? A systematic review on the relative importance of social causation and health selection. Eur J Public Health 2015;25:951–60.

6. Snyder-Mackler N, Sanz J, Kohn JN et al. Social status alters immune regulation and response to infection. Science (80-) 2016;354:1041–5.

7. Sapolsky RM. The influence of social hierarchy on primate health. Science (80-) 2005;308:648–52.

8. Sapolsky RM. Social Status and Health in Humans and Other Animals. Annu Rev Anthropol 2004;33:393–418.

9. Eshetu WB, Woldesenbet SA. Are there particular social determinants of health for the world’s poorest countries? Afr Health Sci 2011;11:108–15.

10. Donkin A, Goldblatt P, Allen J et al. Global action on the social determinants of health. BMJ Glob Heal 2018;3:1–8.

11. Guerra G, Borde E, Salgado de Snyder VN. Measuring health inequities in low and middle income countries for the development of observatories on inequities and social determinants of health. Int J Equity Health 2016;15:9.

12. Henschke N, Mirny A, Haafkens JA et al. Strengthening capacity to research the social determinants of health in low-and middle-income countries: lessons from the INTREC programme. BMC Public Health 2017;17:514.

13. Beehner JC, Bergman TJ. The next step for stress research in primates: To identify relationships between glucocorticoid secretion and fitness. Horm Behav 2017;91:68–83.

14. Simons ND, Tung J. Social Status and Gene Regulation: Conservation and Context Dependence in Primates. Trends Cogn Sci 2019;23:722–5.

15. Anderson JA, Lea AJ, Voyles TN et al. Distinct gene regulatory signatures of dominance rank and social bond strength in wild baboons. Proc R Soc B 2021, DOI: 10.1098/rstb.2020.0441.

16. Jaeggi A V., Blackwell AD, Rueden C von et al. Do wealth and inequality associate with health in a small-scale subsistence society? Elife 2021;10:e59437.

17. Brenner SL, Jones JP, Rutanen-Whaley RH et al. Evolutionary Mismatch and Chronic Psychological Stress. J Evol Med 2015;3:1–11.

18. Gravlee CC. How race becomes biology: Embodiment of social inequality. Am J Phys Anthropol 2009;139:47–57.

19. Lantz PM, House JS, Lepkowski JM et al. Socioeconomic factors, health behaviors, and mortality: results from a nationally representative prospective study of US adults. J Am Med Assoc 1998;279:1703–8.

20. Goldman DP, Smith JP. Can patient self-management help explain the SES health gradient? Proc Natl Acad Sci 2002;99:10929–34.

21. Lynch JW, Kaplan GA, Salonen JT. Why do poor people behave poorly? Variation in adult health behaviours and psychosocial characteristics by stages of the socioeconomic lifecourse. Soc Sci Med 1997;44:809–19.

22. Lahelma E, Lallukka T, Laaksonen M et al. Social class differences in health behaviours among employees from Britain, Finland and Japan: the influence of psychosocial factors. Health Place 2010;16:61–70.

23. Martina Konecna, Urlacher SS. Male social status and its predictors among Garisakang forager-horticulturalists of lowland Papua New Guinea. Evol Hum Behav 2017;8:789–97.

24. Undurraga EA, Nyberg C, Eisenberg DTA et al. Individual wealth rank, community wealth inequality, and self-reported adult poor health: A test of hypotheses with panel data (2002-2006) from native amazonians, bolivia. Med Anthropol Q 2010;24:522–48.

25. Hayward AD, Rickard IJ, Lummaa V. Influence of early-life nutrition on mortality and reproductive success during a subsequent famine in a preindustrial population. Proc … 2013;110:13886–91.

26. Skjærvø GR, Bongard T, Viken Å et al. Wealth, status, and fitness: a historical study of Norwegians in variable environments. Evol Hum Behav 2011;32:305–14.

27. Mealey L. The relationship between social status and biological success: A case study of the Mormon religious hierarchy. Ethol Sociobiol 1985;6:249–57.

28. Clark G, Cummins N. Malthus to modernity: wealth, status, and fertility in England, 1500– 1879. J Popul Econ 2015;28:3–29.

29. Borgerhoff Mulder M, Fazzio I, Irons W et al. Pastoralism and wealth inequality: Revisiting an old question. Curr Anthropol 2010;51:35–48.

30. Johnson BR. Social networks and exchange. In: Little MA, Leslie PW (eds.). Turkana Herders of the Dry Savanna. 1999, 89–106.

31. McCabe JT. Cattle Bring Us to Our Enemies: Turkana Ecology, Politics, and Raiding in a Disequilibrium System. University of Michigan Press, 2004.

32. Mulder MB, Bowles S, Hertz T et al. Intergenerational wealth transmission and the dynamics of inequality in small-scale societies. Science (80-) 2009;326:682–8.

33. Brandolini A, Smeeding T. Income Inequality in Richer and OECD Countries. In: Salverda W, Nolan B, Smeeding T (eds.). The Oxford Handbook of Economic Inequality. Oxford University Press, 2009, 71–100.

34. Sellen DW. Nutritional consequences of wealth differentials in East African pastoralists: The case of the Datoga of northern Tanzania. Hum Ecol 2003;31:529–70.

35. Grandin BE. Wealth and pastoral dairy production: A case study from Maasailand. Hum Ecol 1988;16:1–21.

36. Nestel P. Nutrition of Masaai Women and Children in Relation to Subsistence Food Production. 1985.

37. Paredes Ruvalcaba N, Bignall E, Fujita M. Age and Socioeconomic Status in Relation to Risk of Maternal Anemia among the Ariaal Agropastoralists of Northern Kenya. Hum Ecol 2020;48:47–54.

38. Pike IL, Hilton C, Österle M et al. Low-intensity violence and the social determinants of adolescent health among three East African pastoralist communities. Soc Sci Med 2018;202:117–27.

39. Lea AJ, Martins D, Kamau J et al. Urbanization and market integration have strong, nonlinear effects on cardiometabolic health in the Turkana. Sci Adv 2020;6:eabb1430.

40. Pontzer H, Wood BM, Raichlen DA. Hunter-gatherers as models in public health. Obes Rev 2018;19:24–35.

41. Huneault L, Mathieu MÈ, Tremblay A. Globalization and modernization: An obesogenic combination. Obes Rev 2011;12:64–72.

42. von Rueden CR, Jaeggi A V. Men’s status and reproductive success in 33 nonindustrial societies: Effects of subsistence, marriage system, and reproductive strategy. Proc Natl Acad Sci 2016;113:10824 LP – 10829.

43. Lancaster JB, Kaplan HS. Embodied Capital and Extra-somatic Wealth in Human Evolution and Human History. In: Muehlenbein MP (ed.). Human Evolutionary Biology. Cambridge: Cambridge University Press, 2010, 439–56.

44. Mackinnon DP, Fairchild AJ, Fritz MS. Mediation Analysis. Annu Rev Psychol 2007;58:593–614.

45. Baron R, Kenny D. The Moderator-Mediator Variable Distinction in Social Psychological Research: Conceptual, Strategic, and Statistical Considerations. J Pers Soc Psychol 1986;51:1173–82.

46. Barboza Solís C, Kelly-Irving M, Fantin R et al. Adverse childhood experiences and physiological wear-and-tear in midlife: Findings from the 1958 British birth cohort. Proc Natl Acad Sci U S A 2015;112:E738–46.

47. O’Rand AM, Hamil-Luker J. Processes of Cumulative Adversity: Childhood Disadvantage and Increased Risk of Heart Attack Across the Life Course. Journals Gerontol Ser B 2005;60:S117–24.

48. Shaefer HL, Lapidos A, Wilson R et al. Association of Income and Adversity in Childhood with Adult Health and Well-Being. Soc Serv Rev 2018;92:69–92.

49. Friedman EM, Karlamangla AS, Gruenewald TL et al. Early life adversity and adult biological risk profiles. Psychosom Med 2015;77:176–85.

50. Urlacher SS, Liebert MA, Josh Snodgrass J et al. Heterogeneous effects of market integration on sub-adult body size and nutritional status among the Shuar of Amazonian Ecuador. Ann Hum Biol 2016;43:316–29.

51. McDade T, Nyberg C. Acculturation and health. Human Evolutionary Biology. Cambridge University Press, 2010, 581–602.

52. Adair LS, Popkin BM, Akin JS et al. Cohort profile: The cebu longitudinal health and nutrition survey. Int J Epidemiol 2011;40:619–25.

53. Lamphear J. The People of the Grey Bull: The Origin and Expansion of the Turkana. J Afr Hist 1988;29:27–39.

54. Little MA, Johnson B. Weather conditions in South Turkana, Kenya. In: Dyson-Hudson R, McCabe JT (eds.). South Turkana Nomadism: Coping with an Unpredictably Varying Environment. New Haven: HRAFlex Books, 1985.

55. Galvin K. Nutritional Ecology of Pastoralists in Dry Tropical Africa. Am J Hum Biol 1992;4:209–21.

56. Little MA. Lessons Learned from the South Turkana Ecosystem Project. Hum Ecol Spec Issue No 10 2001:137–49.

57. Felitti VJ, Anda RF, Nordenberg D et al. Relationship of childhood abuse and household dysfunction to many of the leading causes of death in adults. The Adverse Childhood Experiences (ACE) Study. Am J Prev Med 1998;14:245–58.

58. Petruccelli K, Davis J, Berman T. Adverse childhood experiences and associated health outcomes: A systematic review and meta-analysis. Child Abuse Negl 2019;97:104127.

59. Kalmakis KA, Chandler GE. Health consequences of adverse childhood experiences: A systematic review. J Am Assoc Nurse Pract 2015;27:457–65.

60. Hughes K, Bellis MA, Hardcastle KA et al. The effect of multiple adverse childhood experiences on health: a systematic review and meta-analysis. Lancet Public Heal 2017;2:e356–66.

61. Kiburi SK, Molebatsi K, Obondo A et al. Adverse childhood experiences among patients with substance use disorders at a referral psychiatric hospital in Kenya. BMC Psychiatry 2018;18:1–12.

62. Soares ALG, Howe LD, Matijasevich A et al. Adverse childhood experiences: Prevalence and related factors in adolescents of a Brazilian birth cohort. Child Abuse Negl 2016;51:21–30.

63. Meinck F, Cosma AP, Mikton C et al. Psychometric properties of the Adverse Childhood Experiences Abuse Short Form (ACE-ASF) among Romanian high school students. Child Abuse Negl 2017;72:326–37.

64. Mulder MB, Beheim BA. Understanding the nature of wealth and its effects on human fitness. Philos Trans R Soc Lond B Biol Sci 2011;366:344–56.

65. Hruschka DJ, Gerkey D, Hadley C. Estimating the absolute wealth of households. Bull World Health Organ 2015;93:483–90.

66. Sieff DF. The effects of wealth on livestock dynamics among the Datoga pastoralists of Tanzania. Agric Syst 1999;59:1–25.

67. Kaiser BN, Hruschka D, Hadley C. Measuring material wealth in low-income settings: A conceptual and how-to guide. Am J Hum Biol 2017;29:e22987.

68. Novak NL, Allender S, Scarborough P et al. The development and validation of an urbanicity scale in a multi-country study. BMC Public Health 2012;12:530.

69. Cyril S, Oldroyd JC, Renzaho A. Urbanisation, urbanicity, and health: a systematic review of the reliability and validity of urbanicity scales. BMC Public Health 2013;13:513.

70. Liebert MA, Snodgrass JJ, Madimenos FC et al. Implications of market integration for cardiovascular and metabolic health among an indigenous Amazonian Ecuadorian population. Ann Hum Biol 2013;40:228–42.

71. Daly MC, Duncan GJ, McDonough P et al. Optimal indicators of socioeconomic status for health research. Am J Public Health 2002;92:1151–7.

72. Hruschka DJ, Hadley C, Hackman J. Material wealth in 3D: Mapping multiple paths to prosperity in low-and middle-income countries. PLoS One 2017;12:e0184616.

73. “R Core Development Team.” R: A Language and Environment for Statistical Computing. Vienna, Austria: R Foundation for Statistical Computing, 2015.

74. Katsu Y, Iguchi T. Subchapter 95D - Cortisol. In: Takei Y, Ando H, Tsutsui KBT-H of H (eds.). San Diego: Academic Press, 2016, 532–3.

75. Van Cauter E, Leproult R, Kupfer DJ. Effects of gender and age on the levels and circadian rhythmicity of plasma cortisol. J Clin Endocrinol Metab 1996;81:2468–73.

76. Seeman TE, Singer B, Wilkinson CW et al. Gender differences in age-related changes in HPA axis reactivity. Psychoneuroendocrinology 2001;26:225–40.

77. Larsson CA, Gullberg B, Råstam L et al. Salivary cortisol differs with age and sex and shows inverse associations with WHR in Swedish women: a cross-sectional study. BMC Endocr Disord 2009;9:16.

78. Benjamini Y, Hochberg Y. Controlling the false discovery rate: a practical and powerful approach to multiple testing. J R Stat Soc 1995;57:289–300.

79. Lea AJ, Akinyi MY, Nyakundi R et al. Dominance rank-associated gene expression is widespread, sex-specific, and a precursor to high social status in wild male baboons. Proc Natl Acad Sci 2018;115:E12163–71.

80. Narayan KM V, Boyle JP, Thompson TJ et al. Effect of BMI on Lifetime Risk for Diabetes in the U.S. Diabetes Care 2007;30:1562–6.

81. Khan SS, Ning H, Wilkins JT et al. Association of Body Mass Index With Lifetime Risk of Cardiovascular Disease and Compression of Morbidity. JAMA Cardiol 2018;3:280–7.

82. Aune D, Sen A, Prasad M et al. BMI and all cause mortality: systematic review and non-linear dose-response meta-analysis of 230 cohort studies with 3.74 million deaths among 30.3 million participants. BMJ 2016;353, DOI: 10.1136/bmj.i2156.

83. Bynum Griffin, Ridings et al. Adverse childhood experiences reported by adults—Five states, 2009. Morb Mortal Wkly Rep 2010;59:1609–1613.

84. Kamuyango A, Hou W-H, Li C-Y. Trends and Contributing Factors to Contraceptive Use in Kenya: A Large Population-Based Survey 1989 to 2014. Int J Environ Res Public Health 2020;17:7065.

85. Little MA, Gray SJ, Leslie PW. Growth of nomadic and settled Turkana infants of northwest Kenya. Am J Phys Anthropol 1993;92:273–89.

86. Amuna P, Zotor FB. Epidemiological and nutrition transition in developing countries: impact on human health and development: The epidemiological and nutrition transition in developing countries: evolving trends and their impact in public health and human development. Proc Nutr Soc 2008;67:82–90.

87. Slavich GM, Cole SW. The Emerging Field of Human Social Genomics. Clin Psychol Sci 2013;1:331–48.

88. Cole SW. The Conserved Transcriptional Response to Adversity. Curr Opin Behav Sci 2019;28:31–7.

89. Pollitt R, Kaufman JS, Rose KM et al. Early-life and adult socioeconomic status and inflammatory risk markers in adulthood. Eur J Epidemiol 2007;22:55–66.

90. Carroll JE, Cohen S, Marsland AL. Early childhood socioeconomic status is associated with circulating interleukin-6 among mid-life adults. Brain Behav Immun 2011;25:1468–74.

91. Cohen S, Janicki-Deverts D, Chen E et al. Childhood socioeconomic status and adult health. Ann N Y Acad Sci 2010;1186:37–55.

92. Caspi A, Harrington HL, Moffitt TE. Socially Isolated Children 20 Years Later: Risk of Cardiovascular Disease. Arch Pediatr Adolesc Med 2006;160:805–11.

93. Short AK, Baram TZ. Early-life adversity and neurological disease: age-old questions and novel answers. Nat Rev Neurol 2019;15:657–69.

94. Dube SR, Fairweather D, Pearson WS et al. Cumulative childhood stress and autoimmune diseases in adults. Psychosom Med 2009;71:243–50.

95. Kelly-Irving M, Lepage B, Dedieu D et al. Childhood adversity as a risk for cancer: findings from the 1958 British birth cohort study. BMC Public Health 2013;13:767.

96. Kittleson MM, Meoni L a, Wang N-Y et al. Association of childhood socioeconomic status with subsequent coronary heart disease in physicians. Arch Intern Med 2006;166:2356–61.

97. Cluver L, Orkin M, Boyes ME et al. Child and Adolescent Suicide Attempts, Suicidal Behavior, and Adverse Childhood Experiences in South Africa: A Prospective Study. J Adolesc Heal 2015;57:52–9.

98. Anda RF, Butchart A, Felitti VJ et al. Building a Framework for Global Surveillance of the Public Health Implications of Adverse Childhood Experiences. Am J Prev Med 2010;39:93–8.

99. Zefferman MR, Mathew S. Combat stress in a small-scale society suggests divergent evolutionary roots for posttraumatic stress disorder symptoms. Proc Natl Acad Sci 2021;118, DOI: 10.1073/pnas.2020430118.

100. Pike IL, Williams SR. Incorporating psychosocial health into biocultural models: Preliminary findings from Turkana women of Kenya. Am J Hum Biol 2006;18:729–40.

101. Hardt J, Rutter M. Validity of adult retrospective reports of adverse childhood experiences: review of the evidence. J Child Psychol Psychiatry 2004;45:260–73.

102. Favero N, Bullock JB. How (Not) to Solve the Problem: An Evaluation of Scholarly Responses to Common Source Bias. J Public Adm Res Theory 2015;25:285–308.

103. Abbott DH, Keverne EB, Bercovitch FB et al. Are subordinates always stressed? a comparative analysis of rank differences in cortisol levels among primates. Horm Behav 2003;43:67–82.

104. Cole SW. Human Social Genomics. Gibson G (ed.). PLoS Genet 2014;10:e1004601.

105. Borgerhoff Mulder M. Hamilton’s rule and kin competition: the Kipsigis case. Evol Hum Behav 2007;28:299–312.

