## Supplemental Methods and Figures for "Socioeconomic status effects on health vary between rural and urban Turkana"

**Supplementary Materials for: Socioeconomic status effects on health vary between rural and urban Turkana**

The PDF file includes:
Supplementary Methods
Supplementary Figures S1 to S5
Supplementary References

**Supplementary Methods**

**Biomarker measurements**

Body mass index (BMI). Weight was recorded using a portable scale, which was always placed on a hard, flat surface. Height was recorded using a Seca 213 portable stadiometer; before height measurements were recorded, participants were asked to remove any hats or hair ornaments. Weight and height were recorded to the nearest kg and cm, respectively. BMI was calculated as weight (in kg) / height (in m)$^2$.

Waist circumference. Clothing was first removed from the waist line, and the participant was asked to stand with their feet shoulder width apart and their back straight. The top of the hip bone was located by the researcher, and measuring tape was aligned with the top of the hip bone and wrapped around the participant's waist. This procedure was completed three times, and three separate measurements of waist circumference were recorded to the nearest 0.5 cm. We considered the final waist circumference value for a given participant to be the average of the three measurements.

Lipid profiles. Approximately 8mL of intravenous blood was collected into an EDTA vacutainer tube. Immediately after collection, 40ul of each sample was used to conduct a lipid panel using the CardioCheck Plus (PTS Diagnostics) following the manufacturer's instructions. Specifically, total cholesterol, HDL cholesterol, and triglycerides were measured for each participant, and LDL cholesterol was calculated by the CardioCheck Plus. Because the CardioChek Plus cannot read triglyceride values below 50 mg/dL, both triglyceride and LDL cholesterol values were not recorded for these individuals; this is because LDL cholesterol is estimated as a function of triglyceride, HDL cholesterol, and total cholesterol levels using the Friedewald Equation (LDL cholesterol = total cholesterol - HDL cholesterol – triglycerides/5).

Blood glucose levels. A drop of whole blood (~10ul) from a finger prick using a safety lancet was used to measure glucose levels. To do so, we used the OneTouch system following the manufacturer's instructions.

Body fat percentage. Body fat percentage was measured via bioelectrical impedance using the Omron HBF-306C Handheld Body Fat Loss Monitor according to the manufacturer's instructions.

Blood pressure. Arterial blood pressure was measured as systolic blood pressure and diastolic blood pressure. All participants sat in a relaxed position with their arm flat at a 90-degree angle, and with the arm relaxed and the wrist facing up. The cuff was placed around the participant's upper arm (approximately ½ inch above the elbow). Participants were asked not to speak while a trained assistant used an Omron 10 Series Wireless Upper Arm Blood Pressure Monitor to collect a single measurement.

**Analyses using alternative measures of SES**

We performed two supplementary analyses using measures of wealth that were directly comparable across contexts. First, we used the measures of wealth described in the main text for pastoralists (livestock holdings) and for urban individuals (tally of durables/goods, dwelling characteristics, and other household assets), but applied these measures to the analysis of health outcomes in both contexts. The full results from these analyses are presented in Tables S1 and S12. We find that a tally of assets does not predict any of our cardiometabolic health or self-reported health measures in the pastoralist setting (all p>0.05), probably because there is very little variation in this measure (3.1% of individuals have non-zero values). In contrast, greater livestock holdings predict worse health in the urban setting, namely higher triglycerides (β SES=0.056, q-value=0.033). We also observed marginally significant results indicating that greater livestock holdings predict higher cholesterol levels (β SES=0.053, p-value=0.050), higher blood glucose levels (β SES=0.047, p-value=0.045), higher BMI (β SES=0.045, p-value=0.034), and greater incidence of fatigue or weakness (β SES=0.453, p-value=0.015). As noted in the main text, livestock holdings are correlated with the assets tally among urban individuals (β=0.027, p-value=0.036, Poisson regression), which explains why these measures are both associated worse health outcomes in this context.

In a second analysis, we followed the logic of [1,2] and performed principal components analysis on the following variables, all of which are potential indicators of socioeconomic status: highest education level (coded as a numeric variable ranging from 0-4), finished floor, finished roof, electricity, television set, mobile phone, flush toilet, treated water, indoor tap water, gas cooking, total livestock holdings (log2 transformed), ratio of household members to number of rooms in the house. The first principal component (PC 1) explained 59.34% of the variance and was very strongly predicted by lifestyle (linear model predicting PC 1 as a function of lifestyle, β=-5.528, p-value<$10^{-10}$). PC 1 was negatively associated with highest education level (Pearson correlation, R=-0.547) as well as ownership of a finished floor (R=-0.436), a finished roof (R=-0.636), electricity (R=-0.423), a television set (R=-0.318), a mobile phone (R=-0.527), a flush toilet (R=-0.203), methods for treating water (R=-0.343), an indoor tap (R=-0.235), and methods for gas cooking (R=-0.294); it was also positively associated with livestock holdings (R=0.907) and the ratio of household members to number of rooms in the house (R=0.675). We found that higher PC 1 values predicted better health among pastoralists, namely lower incidence of chest pain (β SES=-0.578, q-value= 0.017), fatigue or weakness (β SES=-0.788, q-value= 0.058) and marginally lower incidence of diarrhea (β SES=-0.850, p-value=0.033). Higher PC 1 values predicted worse health among urban individuals, though only at a nominal p-value cutoff of 0.05: specifically, greater incidence of chest pain (β SES=0.443, p-value=0.019). The full results from these analyses are presented in Table S13. Overall, while power is reduced by using a measure that is not tailored to a given context, the direction of the SES-health associations we observe here is entirely consistent with the results presented in the main text.

**Supplementary Figures**

**Supplementary Figure 1. Sampling locations.** Each dot represents an individual sample, points are jittered for visualization. The major urban centers included in this study, namely Lodwar, Kitale, and Nanyuki, are marked with a black dot.

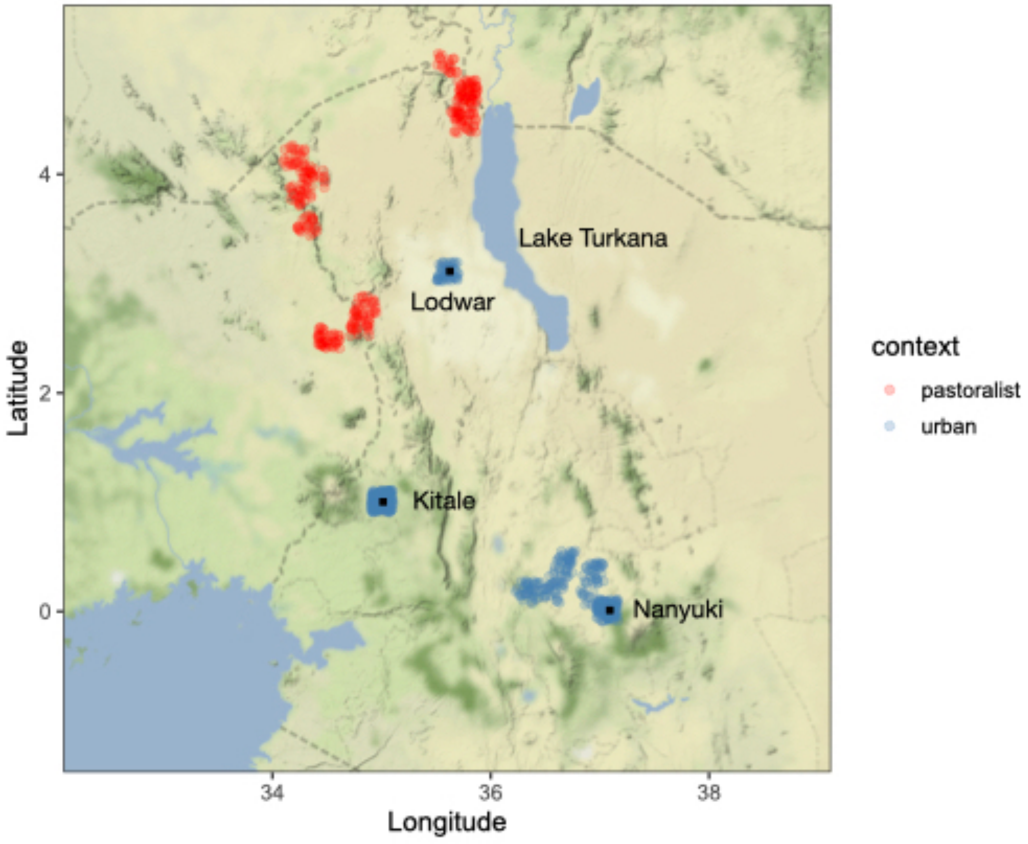

**Supplementary Figure 2. Distribution of cardiometabolic biomarkers among pastoralist and urban Turkana.** Dots represent the median of each distribution and lines represent the median +/- 1 standard deviation. Values are broken down by sex for each group (F=female, M=male).

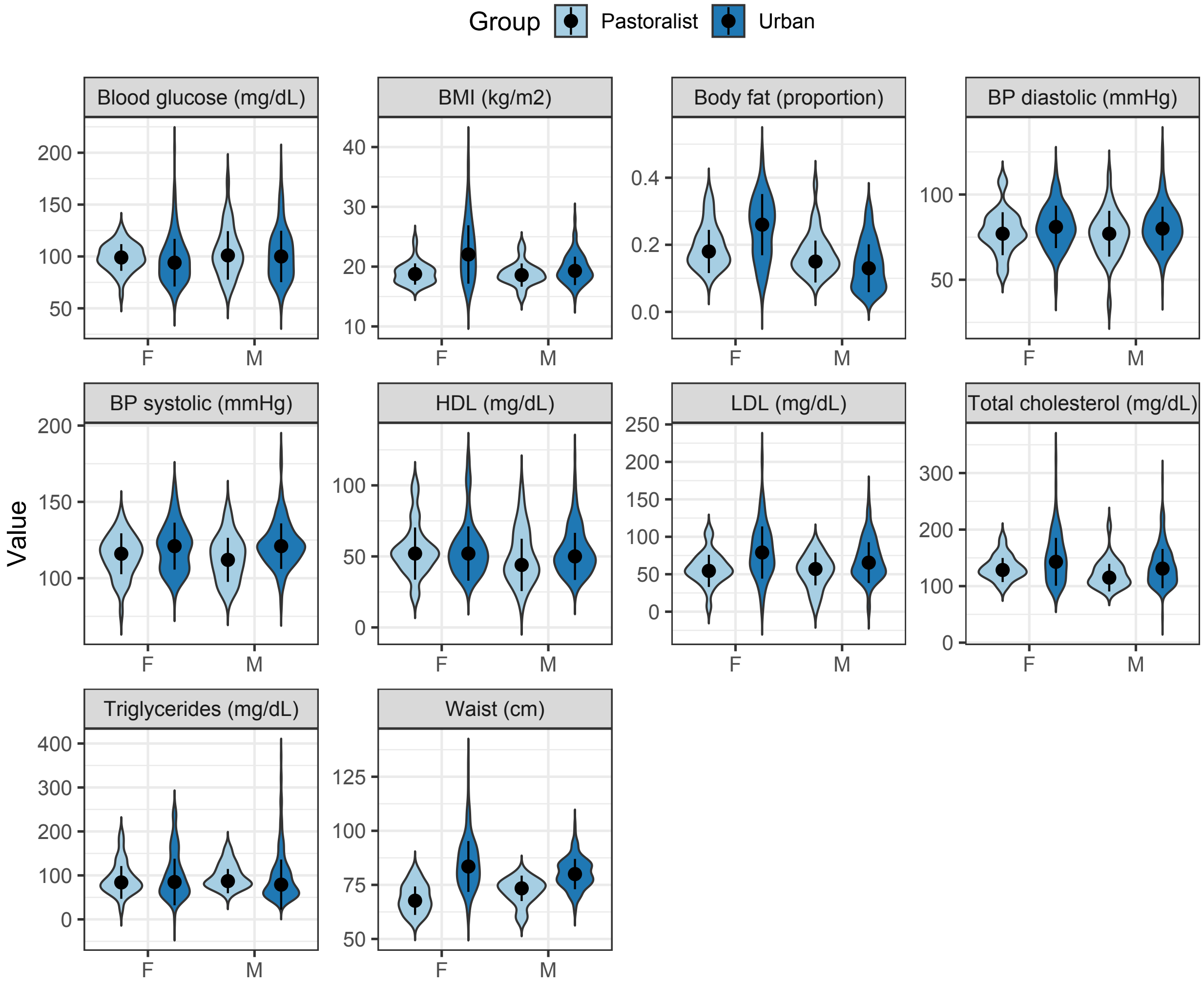

**Supplementary Figure 3. Among pastoralist men, high socioeconomic status predicts more wives (A), which predicts more children (B).** In panel A, data are plotted for individuals in from the highest and lowest SES quartiles for visualization. In panel B, data from 5 individuals with >3 wives were removed.

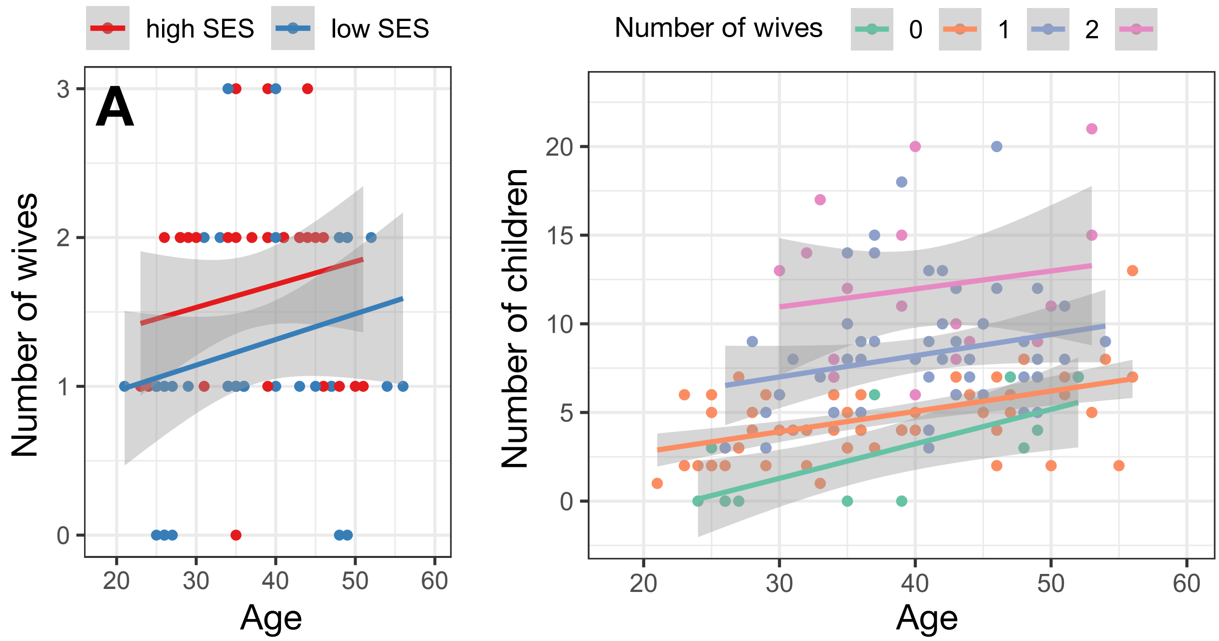

**Supplementary Figure 4. Cortisol results.** Bivariate relationships between cortisol and (A) sex, (B) age, and (C) time of day in the total sample (n=216). Bivariate relationships between cortisol and SES among pastoralist (panel D; n=79) and urban (panel E; n=59) individuals, respectively. In panel A, blue represents males and pink represents females.

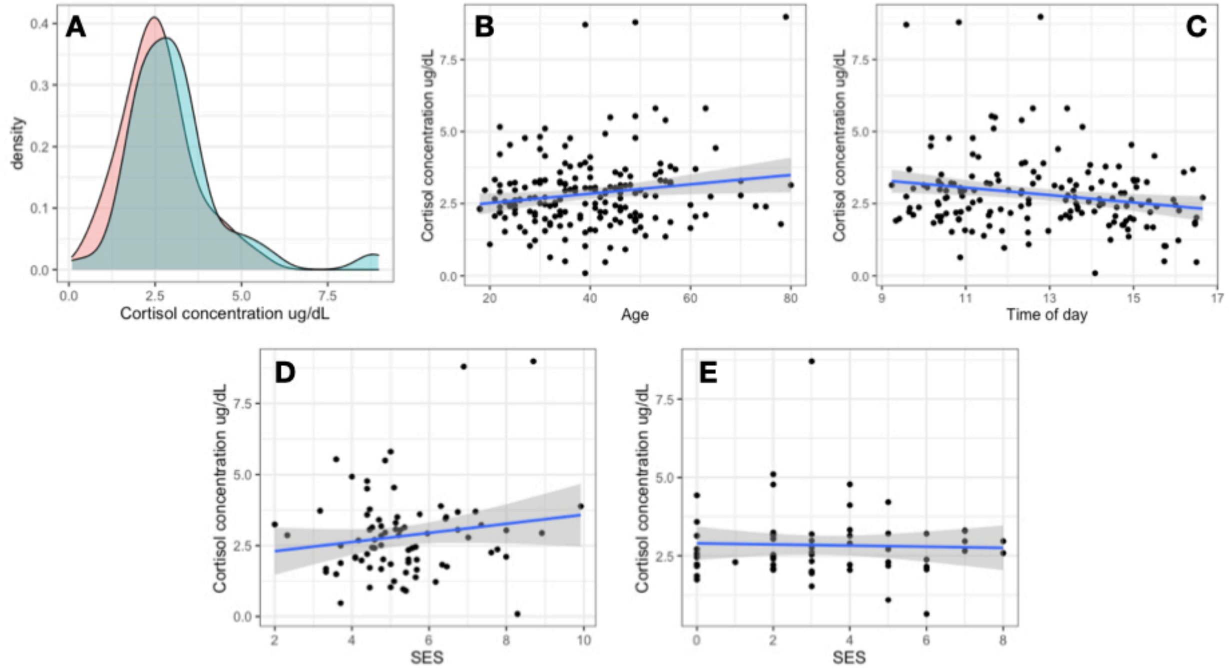

**Supplementary Figure 5. Proportion of individuals experiencing specific early life adversities among urban Turkana, pastoralist Turkana, and US individuals.** US data were sourced from [3].

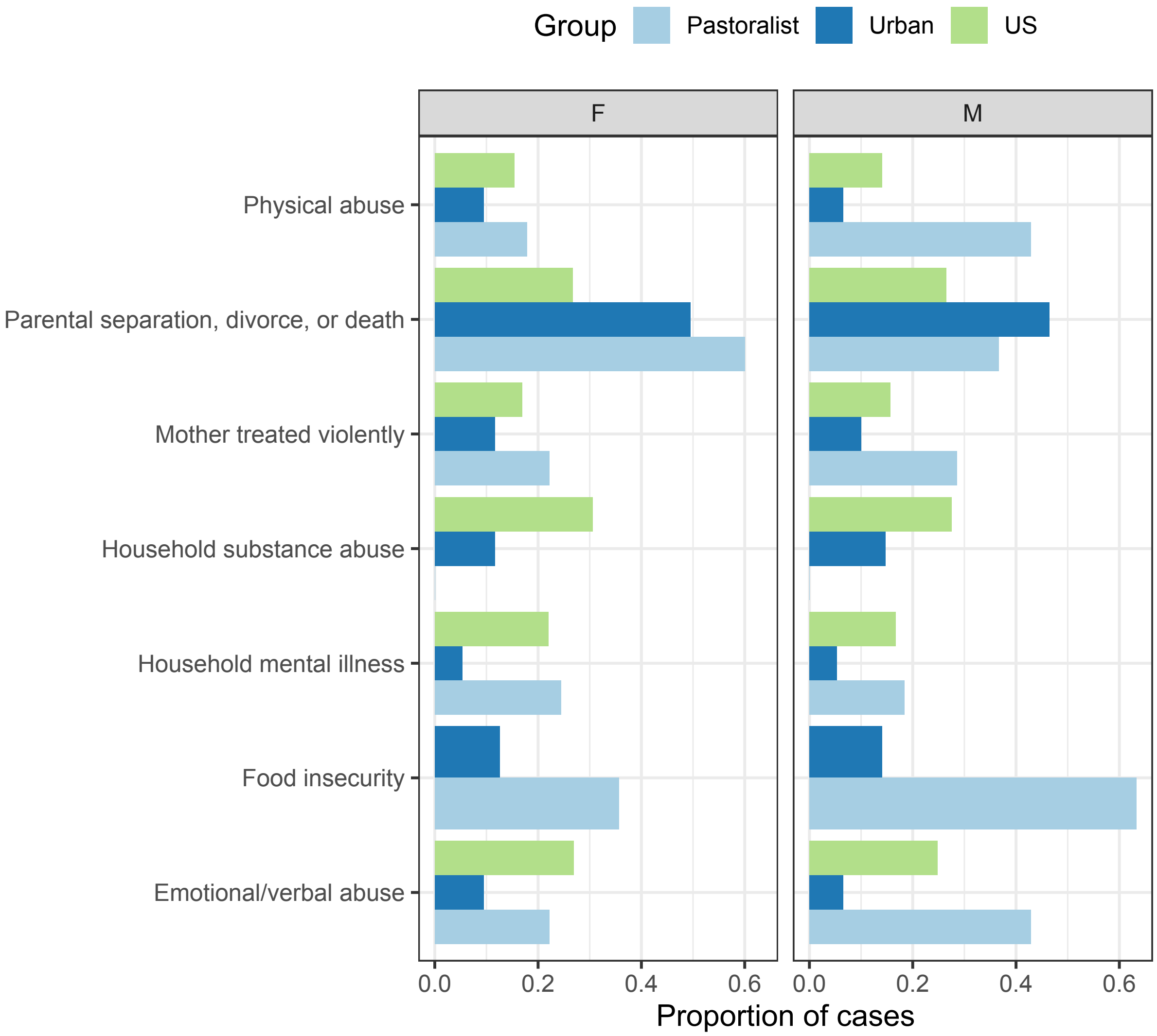

**Supplementary References**

1. Hruschka DJ, Hadley C, Hackman J. Material wealth in 3D: Mapping multiple paths to prosperity in low- and middle- income countries. *PLoS One* 2017;**12**:e0184616.
2. Vyas S, Kumaranayake L. Constructing socio-economic status indices: how to use principal components analysis. *Health Policy Plan* 2006;**21**:459–68.
3. Bynum, Griffin, Ridings *et al.* Adverse childhood experiences reported by adults—Five states, 2009. *Morb Mortal Wkly Rep* 2010;**59**:1609–1613.
